# Investigating associations between blood metabolites, later life brain imaging measures, and genetic risk for Alzheimer’s disease

**DOI:** 10.1101/2022.01.29.22270098

**Authors:** Rebecca Green, Jodie Lord, Marzia A. Scelsi, Jin Xu, Andrew Wong, Sarah Naomi-James, Alex Handy, Lachlan Gilchrist, Dylan M. Williams, Thomas D. Parker, Christopher A. Lane, Ian B. Malone, David M. Cash, Carole H. Sudre, William Coath, David L. Thomas, Sarah Keuss, Richard Dobson, Cristina Legido-Quigley, Nick C. Fox, Jonathan M. Schott, Marcus Richards, Petroula Proitsi, The Insight 46 study team

## Abstract

Identifying blood-based signatures of brain health and preclinical pathology may offer insights into early disease mechanisms and highlight avenues for intervention. Here, we systematically profiled associations between blood metabolites and whole-brain volume, hippocampal volume, and amyloid-β status among participants of Insight 46 – the neuroscience sub-study of the National Survey of Health and Development (NSHD). We additionally explored whether key metabolites were associated with polygenic risk for Alzheimer’s disease (AD).

Following quality control, concentrations of 1019 metabolites – detected with liquid chromatography-mass spectrometry – were available for 1740 participants at age 60-64. Metabolite data were subsequently clustered into modules of co-expressed metabolites using weighted coexpression network analysis. Accompanying MRI and amyloid-PET imaging data were present for 437 participants (age 69-71). Regression analyses tested relationships between metabolite measures – modules and hub metabolites – and imaging outcomes. Hub metabolites were defined as metabolites that were highly connected within significant (p_FDR_<0.05) modules or previously identified as a hub for cognition in the same cohort. Regression models included adjustments for age, sex, *APOE* genotype, lipid medication use, childhood cognition and social factors. Finally, AD polygenic risk scores (PRS), including and excluding the *APOE* region, were tested for relationships with metabolites and modules that associated (p_FDR_<0.05) with an imaging outcome (N=1638).

In the fully adjusted model, three lipid modules were associated with a brain volume measure (p_FDR_<0.05): one enriched in sphingolipids (hippocampal volume: ß=0.14, 95%CI=[0.055,0.23]), one in several fatty acid pathways (whole-brain volume: ß=-0.072, 95%CI=[-0.12,-0.026]), and another in diacylglycerols and phosphatidylethanolamines (whole-brain volume: ß=-0.066, 95%CI=[-0.11,-0.020]). Twenty-two hub metabolites were associated (p_FDR_<0.05) with an imaging outcome (whole-brain volume: 22; hippocampal volume: 4). Some nominal associations were reported for amyloid-β, and with an AD PRS in our genetic analysis, but none survived multiple testing correction.

Our findings highlight key metabolites, with functions in membrane integrity and cell signalling, that associated with structural brain measures in later life. Future research should focus on replicating this work and interrogating causality.

## Background

Brain changes accompanying ageing are varied and can include pathologies that lead to cognitive impairment, the commonest of which is Alzheimer’s disease (AD). Identifying non-invasive and scalable markers of brain health and pathology in later life, including but not limited to those associated with AD, would be valuable for research and therapeutic trials. This has led to large efforts in detecting blood-based markers, with candidates such as neurofilament light and phosphorylated-tau showing particular promise ^1^. Blood metabolites – the products of chemical reactions occurring in the body – may also present as potential candidates for this goal. Due to their proximity to core biological processes, they are uniquely placed to capture real-time physiological changes and may allow insights into the processes associated with emergence of disease ^2^. Additionally, since they are potentially modifiable ^3,4^, they could represent possible targets for intervention.

Existing research has identified associations between several metabolite classes and imaging markers related to neurodegeneration, including particular lipids and amino acids ^5–9^. However, these studies have been directed towards clinical cohorts and thus may not be representative of the general preclinical population. Additionally, little is known about the involvement of groups of interrelated metabolites; utilising systems-level approaches could offer an improved understanding of these complex relationships and facilitate the identification of candidate markers. We previously employed a systems-level approach to explore the metabolic correlates of late midlife cognition in the Medical Research Council National Survey of Health and Development (NSHD; the British 1946 birth cohort) ^10^. We identified groups of highly coexpressed metabolites that associated with cognitive outcomes and key metabolites within these to explore further, including acylcarnitines, modified nucleotides and amino acids, vitamins, and sphingolipids; although many associations with late midlife cognition were explained by social factors and childhood cognition ^10^.

Here, we aimed to investigate the metabolic correlates of later life brain imaging measures relevant to AD and neurodegeneration – Aβ pathology, whole-brain volume and hippocampal volume. To provide a deeper understanding on the nature of potential relationships and how they may contribute to AD risk, we explored whether any key metabolites were additionally associated with polygenic risk for AD.

## Methods

### Participants

The NSHD is a broadly representative birth cohort study, originally following 5362 individuals since their birth in mainland Britain during one week in March 1946 ^11^. At age 69-71, 502 participants enrolled in Insight 46, the neuroscience sub-study. At a University College London clinic they underwent comprehensive clinical and cognitive tests, MRI, and ^18^F-florbetapir positron emission tomography (PET) imaging ^12,13^. Compared to the full NSHD cohort, participants of Insight 46 were of slightly higher cognitive ability, more socially advantaged, and of better overall health ^13^. Further details on participant eligibility and recruitment can be found elsewhere ^12^.

Ethical approval was obtained from the National Research Ethics Service Committee London (14/LO/1173). All participants provided written informed consent.

## Materials

### Metabolite quality control

At age 60-64, blood samples were collected by trained research nurses. Samples were stored at - 80 degrees.

Using Ultrahigh Performance Liquid Chromatography-Tandem Mass Spectrometry (UPLC-MS/MS), concentrations of 1401 metabolites were detected and measured by Metabolon Inc (Durham, NC, USA) among 1814 NSHD participants. Metabolites were assigned to nine families (lipids, amino acids, xenobiotics, peptides, nucleotides, cofactors and vitamins, carbohydrates, energy and partially characterised molecules) and further organised into pathways based on their proposed biological function informed by the Kyoto Encyclopaedia of Genes and Genomes (KEGG) database (Supplementary Table 1). Unknown metabolites were assigned to an “Unknown” family and pathway and denoted by a number prefixed by an “X”; these were included in all analyses.

Metabolite data underwent strict quality control (QC), as detailed in ^10^, resulting in 1019 metabolites (Supplementary Notes).

### Genetic quality control

Initial QC and imputation were performed centrally by the NSHD study team (Supplementary Notes). For this analysis, we removed variants that were rare (MAF <5%), with a low call rate (<98%), or that deviated from Hardy-Weinberg equilibrium (p <1×10^−5^). We additionally removed participants with a low call rate (<98%), mismatching biological and reported sex, or that were related (PIHAT <0.1). All QC was performed using PLINK v1.9 (https://www.cog-genomics.org/plink2) ^14^. Following QC, genetic and metabolomic data were available for 1638 participants.

### Scanning procedure

The scanning procedure and data processing were undertaken by the Insight 46 team. Aβ-PET and MRI were acquired contemporaneously using a single Biograph mMR 3 Tesla PET/MRI scanner (Siemens Healthcare) ^12^. Aβ burden was quantified over 10 minutes, approximately 50 minutes after intravenous injection with ^18^F-florbetapir (370 mBq. Standardised uptake value ratios (SUVRs) were derived using a grey matter cortical composite and an eroded subcortical white matter reference region. A cut-off of >0.6104 was used to define Aβ positivity being the 99th percentile of the lower (Aβ-negative) Gaussian distribution ^15^. Participants below this threshold were defined as Aβ-negative. Data were processed using an in-house pipeline, including attenuation correction using pseudo-CT ^12^. For volumetric T1-weighted MRI images, visual QC was performed as detailed in ^12^, and processed using the following: MAPS ^16^ for whole-brain volume (with manual editing if needed); STEPS ^17^ for left and right hippocampal volumes (with manual editing if needed); and SPM12 (<u>fil.ion.ucl.ac.uk/spm</u>) ^18^ for total intracranial volume (TIV). Hippocampal volume was calculated as the mean volume of the left and right hippocampi.

### Covariables

In line with our previous analysis in the full NSHD ^10,19^, covariables were: sex, blood collection information (clinic location, age, fasting status), age at scan, *APOE* genotype (ε4 carrier/non carrier; blood samples at age 53 or 69y), BMI (60-64y nurse visit), lipid medication (yes/no; self-reported use in 24 hours preceding blood collection at 60-64y), childhood cognition (15y), highest level of educational attainment (no qualifications/’O level’/’A level’ or higher; 26y), childhood socioeconomic position (SEP) (Father’s current or last known occupation categorised according to the UK Registrar General; 11y), and midlife SEP (own occupation categorised as for childhood SEP; 53y).

### Statistical analysis

We previously imputed missing covariable data using multiple imputation chained equations (100 iterations and 50 imputations) ^20^ in the full NSHD metabolomics dataset. Further details of missing data can be found in Table 1. Unless otherwise specified, we carried out all analyses in R version 3.6.0 (details of all software and packages used can be found in Supplementary Notes). A visual summary of our analytical pipeline can be found in Figure 1.

**Table 1.** Characteristics of participants included in this analysis.

|  |  | Participants with genetic and metabolite data |  | Participants with imaging and metabolite data |  |
| --- | --- | --- | --- | --- | --- |
|  |  | Overall | Missing (%) | Overall | Missing (%) |
| **n** |  | 1638 |  | 437 |  |
| **Sex, N (%)** | Male | 812 (49.6) | 0 | 229 (52.4) | 0 |
|  | Female | 826 (50.4) |  | 208 (47.6) |  |
| **Age at scan (years), mean (SD)** |  |  |  | 70.7 (0.7) | 0 |
| **APOE4 carrier, N (%)** | Non-carrier | 1044 (69.6) | 8.5 | 306 (70.3) | 0.5 |
|  | Carrier | 455 (30.4) |  | 129 (29.7) |  |
| **Amyloid status, N (%)** | Negative |  |  | 348 (81.1) | 1.8 |
|  | Positive |  |  | 81 (18.9) |  |
| **Hippocampal volume in ml, mean (SD)** |  |  |  | 3.1 (0.3) | 0.5 |
| **Brain volume in ml, mean (SD)** |  |  |  | 1101.8 (99.3) | 0.5 |
| **Total intracranial volume in ml, mean (SD)** |  |  |  | 1435.9 (132.2) | 0.5 |
| **Age at blood collection (years), mean (SD)** |  | 63.2 (1.1) | 0 | 63.3 (1.1) | 0 |
| **Fasting at blood collection , N (%)** | No | 63 (3.8) | 0 | 6 (1.4) | 0 |
|  | Yes | 1575 (96.2) |  | 431 (98.6) |  |
| **Lipid medication use (age 60-64), N (%)** | No | 1232 (75.2) | 0 | 335 (76.7) | 0 |
|  | Yes | 406 (24.8) |  | 102 (23.3) |  |
| **Body mass index (age 60-64) in kg/m2, mean (SD)** |  | 27.8 (4.7) | 0.1 | 27.4 (4.0) | 0 |
| **Childhood cognitive ability (age 15)\*, z-score, mean (SD)** |  | 0.2 (0.8) | 14.3 | 0.5 (0.7) | 8.2 |
| **Childhood socioeconomic position (age 11), N (%)** | Unskilled | 80 (5.1) | 5 | 18 (4.1) | 0.5 |
|  | Partly skilled | 274 (17.6) |  | 64 (14.7) |  |
|  | Manual skilled | 462 (29.7) |  | 106 (24.4) |  |
|  | Nonmanual skilled | 276 (17.7) |  | 90 (20.7) |  |
|  | Intermediate | 346 (22.2) |  | 110 (25.3) |  |
|  | Professional | 118 (7.6) |  | 47 (10.8) |  |
| **Adult socioeconomic position (age 53), N (%)** | Unskilled | 46 (2.8) | 0.4 | 4 (0.9) | 0 |
|  | Partly skilled | 159 (9.7) |  | 22 (5.0) |  |
|  | Manual skilled | 238 (14.6) |  | 40 (9.2) |  |
|  | Nonmanual skilled | 380 (23.3) |  | 91 (20.8) |  |
|  | Intermediate | 680 (41.7) |  | 225 (51.5) |  |
|  | Professional | 129 (7.9) |  | 55 (12.6) |  |
| **Highest educational attainment (age 26), N (%)** | No qualification | 446 (28.6) | 4.9 | 65 (15.3) | 3 |
|  | Up to GCSE | 446 (28.6) |  | 125 (29.5) |  |
|  | A-level or higher | 665 (42.7) |  | 234 (55.2) |  |

\*Childhood cognition Z-scores were calculated in the full National Survey of Health and Development cohort (N=5362)

**Figure 1.**
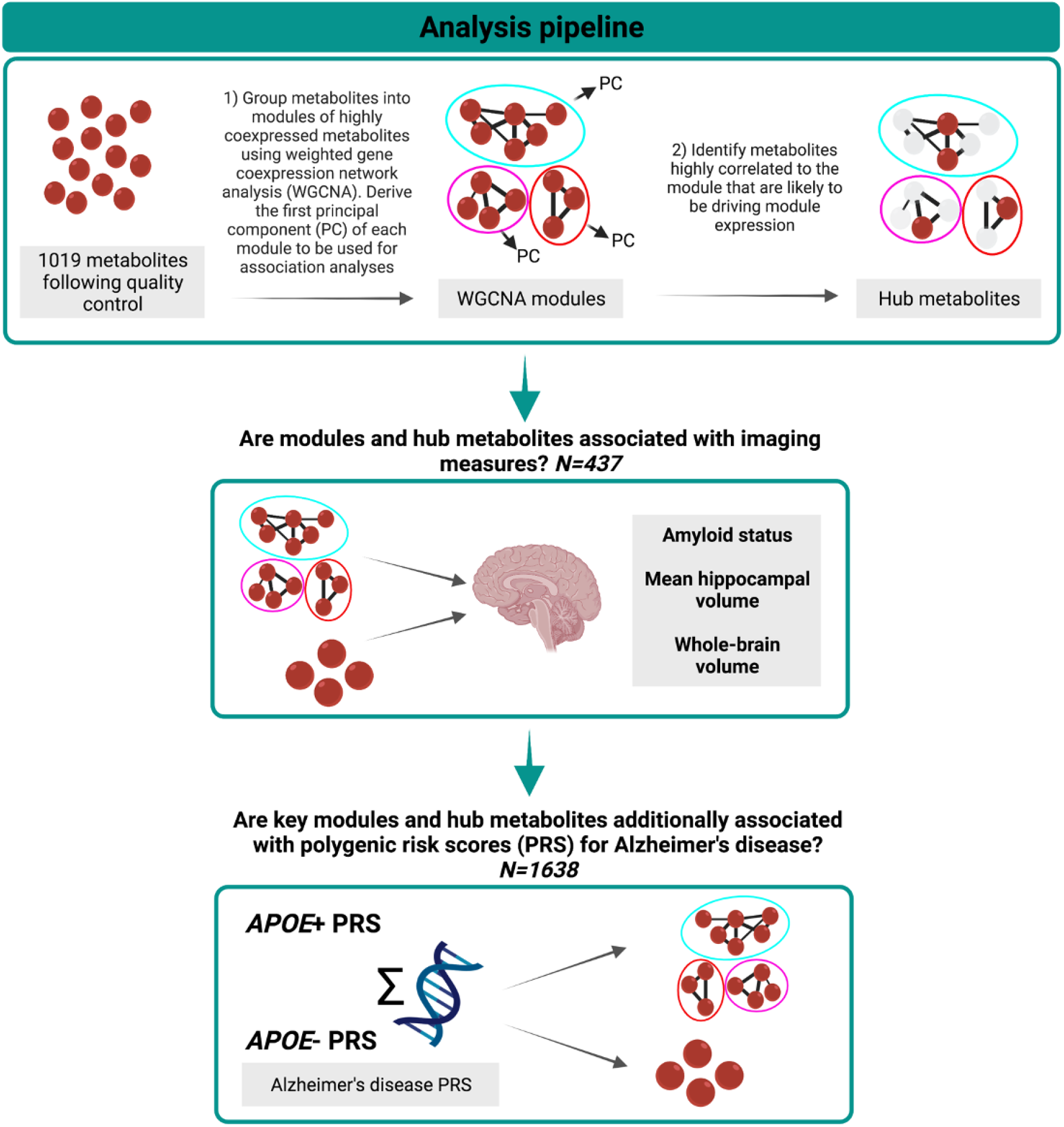
Analysis pipeline.

### Coexpression network analysis

To explore associations of clusters (termed “modules”) of co-expressed metabolites, we applied weighted gene coexpression network analysis (WGCNA) ^21–23^ to metabolite data, as detailed previously ^10^. First, metabolite data were adjusted for sex and blood clinic information and the standardised residuals were used for WGCNA. Fourteen modules of highly connected metabolites were then identified, and the first principal component of each module (termed “module eigenvalue”) was derived to allow for relationships between modules and outcomes to be examined. Overrepresentation analyses were conducted, using the hypergeometric test, to identify enriched pathways within the module and provide insight into potential biological function ^10^. Modules were allocated an arbitrary colour name using the WGCNA package for ease of discussion..

### Hub metabolites

Metabolites that are highly connected to their module (termed “hub metabolites”) are likely to be functionally important and thus present as valuable marker candidates ^24^. We previously identified associations between 35 hubs, defined using correlations between metabolites and module eigenvalues exceeding r=0.65 (termed “module membership”; kME), and cognitive outcomes in the NSHD ^10^. As these metabolites were selected on the premise of showing associations with cognition, we additionally looked for hubs that may be important in brain imaging outcomes. To do this, we extracted any additional metabolites exceeding the 0.65 threshold ^10^ in modules showing significant (p_FDR_<0.05) associations in the present analysis.

### Regression analysis

To allow for direct comparisons, we standardised all continuous predictors and outcomes to a mean of 0 and standard deviation of 1. We then tested relationships between a) modules, and b) hub metabolites using linear regression (for whole-brain volume and hippocampal volume) and logistic regression (for Aβ status). Model 1 adjusted for basic covariables: sex, blood collection information, age at scan, *APOE* genotype, and total intracranial volume (for whole-brain volume and hippocampal volume only). As modules were already adjusted for sex and blood clinic information, these covariables were not additionally included for module analyses. Model 2 additionally adjusted for lipid-related factors: BMI and lipid medication use. Finally, model 3 further adjusted for childhood cognition, educational attainment, and SEP (parental and midlife).

Analyses were conducted on each imputed dataset and pooled using Rubin’s rules ^25^. Regression assumptions were checked by examination of the residuals. We applied False Discovery Rate (FDR) correction using the Benjamini-Hochberg procedure ^26^ with an alpha=0.05. FDR correction was applied separately to each analysis (module and hub) and each outcome.

### Polygenic risk scores

To explore whether levels of key metabolites were influenced by genetic risk for AD, we investigated associations between AD polygenic risk scores (PRS) – a weighted sum of genetic variants associated with a trait or disease – and significant (p_FDR_<0.05) hub metabolites and modules. We obtained genome-wide association study summary statistics from Kunkle et al. ^27^ (N=63,926; 21,982 AD clinically-ascertained cases, 41,944 controls), which were used as the base data for PRS analyses. Using PRSice-2 ^28^, we computed PRS in the NSHD, both including and excluding SNPs in the *APOE* region (chr 19, GRCh37 coordinates 44912079 to 45912079) ^29^. Two p-value thresholds (P_T_) – previously identified to be optimal for PRS including and excluding the *APOE* region – were used for SNP selection: 5×10^−8^ (suggested for *APOE* region included) and 0.1 (suggested for *APOE* region excluded) ^30^, resulting in four PRS. SNPs in linkage disequilibrium (r^2^>0.001 within a 250kb window) were clumped and the SNP with the lowest p-value was retained.

We first standardised predictors and outcomes to a mean of 0 and standard deviation of 1. Then, we regressed PRS on key metabolites and modules, adjusting for sex, age, blood collection details, and seven principal components (to control for population stratification). We applied FDR correction to each analysis – module and hub metabolite – using the Benjamini-Hochberg procedure ^26^ with an alpha=0.05.

### Additional analysis

We conducted several additional analyses to test the robustness of our findings (see Supplementary Notes for full details). In brief, we first investigated whether WGCNA modules, which were curated in the full NSHD, were preserved in the Insight 46 subset. Then, for our main analyses, we applied a more conservative Bonferroni correction to our findings (module: p<3.57×10^−3^; hub metabolite: p<1.06×10^−3^). Finally, we explored whether lifestyle and related factors (lifetime smoking, diet, exercise, blood pressure, and alcohol intake) explained any of our results.

## Results

### Participant characteristics

Insight 46 participants who completed the scanning procedure, were dementia-free, and with full metabolite data were included for module and hub metabolite analysis (N=437; 47.6% female, 18.9% Aβ-positive). For PRS analysis, NSHD participants with metabolomics and genetic data were included (N=1638; 50.4% female). Participant characteristics can be found in Table 1 (see Supplementary Notes for characteristics split by Aβ status).

### Metabolite coexpression network modules

Overall, we identified three modules that showed associations with brain volume outcomes (p_FDR_<0.05) and none (p>0.05) with Aβ status. Full results can be found in Supplementary Table 2 and are visualised in Figure 2. Results of the fully adjusted model are discussed hereafter.

**Figure 2.**
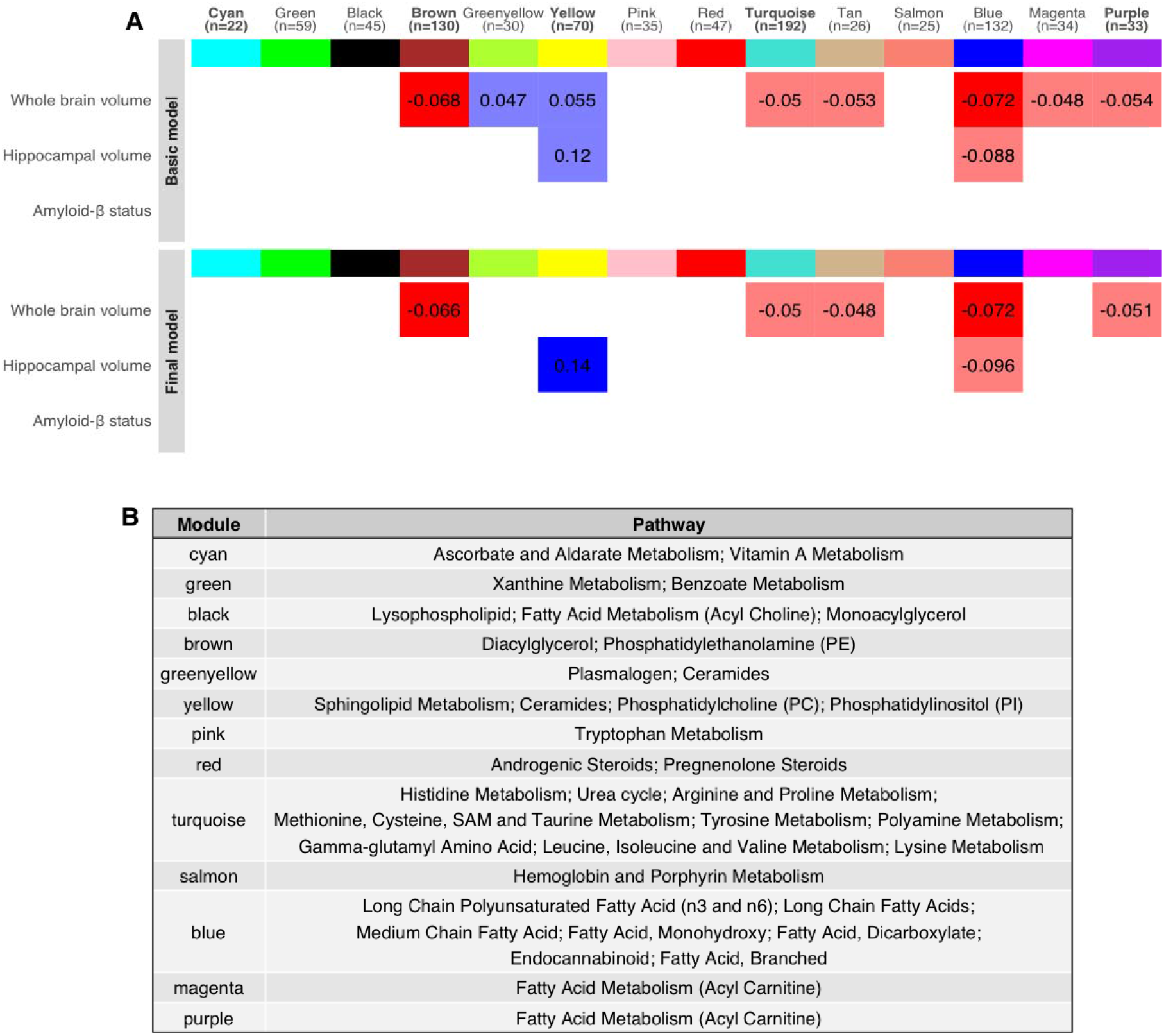
Module results. A) Heatmap showing relationships between modules and brain imaging outcomes in the basic model and final model. Tiles are coloured by effect direction (blue = associated with better outcomes, red = associated with worse outcomes). Effect sizes are presented inside the tiles. Associations significant after multiple testing correction are represented by a solid fill and nominal (p<0.05) by a fainter fill. Module names in bold were additionally associated with a cognitive outcome in our previous analysis. Source data are present in Supplementary Table 2. B) Table presenting enriched pathways in each module, with the most highly enriched pathways presented first. No pathways were enriched for the tan module. Source data are available in ^10^

We report associations between higher expression of two lipid modules and smaller whole-brain volumes: the brown module, enriched in diacylglcerol (DAG) and phosphatidylethanolamine (PE) pathways (ß=-0.066, 95%CI=[-0.11,-0.019], p=0.006, p_FDR_=0.044) and the blue module, enriched in various fatty acids pathways (ß =-0.072, 95%CI=[-0.12,-0.026], p=0.0021, p_FDR_=0.035). Higher expression of the yellow module, enriched in sphingolipid metabolism and related pathways, was associated with a larger hippocampal volume (ß =0.14, 95%CI=[0.055,0.23], p=0.0017, p_FDR_=0.035).

### Hub metabolites

We explored associations between 81 metabolites that were highly connected (kME>0.65) in significant modules identified in 3.2, and 35 that were identified to be hubs in our previous study on cognition. Across all models, we report 30 key metabolites after FDR correction, of which 13 were previously associated with cognitive outcomes (Figure 3). No associations were detected for any metabolite and Aβ status after FDR correction, although some nominal associations were observed (3 cyan and 1 yellow metabolite; Supplementary Table 1 & Figure 3). In the fully adjusted model, 22 metabolites were associated (pFDR<0.05) with an imaging outcome (whole-brain volume: 22; hippocampal volume only: 4). Of the 22, 10 metabolites belonged to the yellow module (pathways: sphingolipid metabolism) and were positively associated with larger brain volumes. Twelve metabolites were negatively associated with brain volumes: six belonging to the blue module (pathways: fatty acid (monohydroxy; dicarboxylate), long chain PUFA (n3 and n6), glycerolipid metabolism), four to the brown module (pathways: PE, phosphatidylcholine (PC), DAG), and two to additional modules identified in our previous analysis of cognition (pathways: fatty acid metabolism (acyl carnitine); methionine, cysteine, SAM and taurine metabolism).

**Figure 3.**
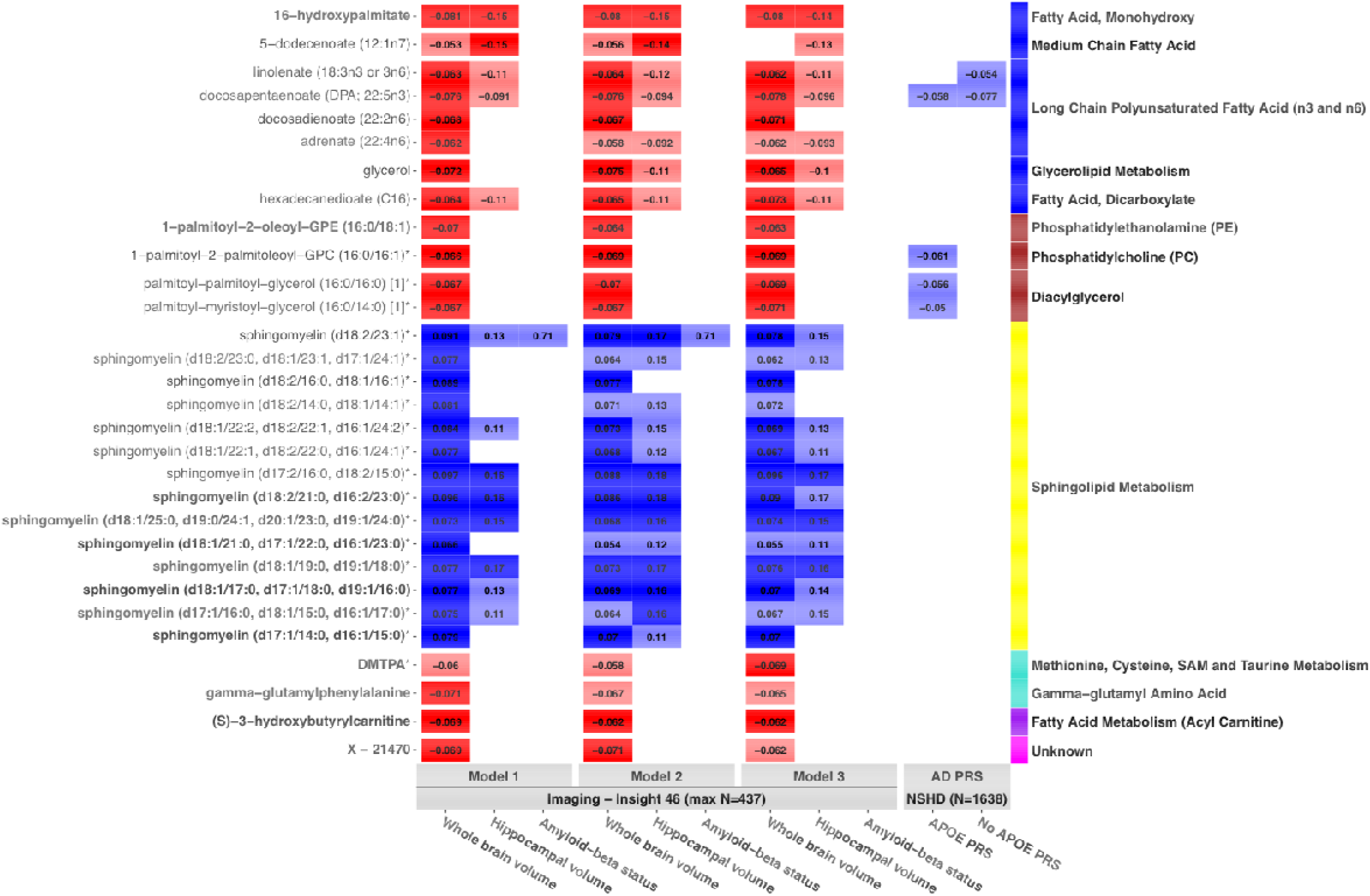
Hub metabolite results. Heatmap showing relationships between key hub metabolites (pFDR<0.05) and brain imaging outcomes, as well as relationships between thes e metabolites and Alzheimer’s disease polygenic risk scores (best threshold shown). Tiles are coloured by effect direction (blue = associated with better outcomes, red = associated with worse outcomes). Effect sizes are presented inside the tiles. Associations significant after multiple testing correction (pFDR<0.05) are represented by a solid fill and nominal (p<0.05) by a fainter fill. Metabolites are organised by both module (indicated via the colour panel on the right) and pathway (specified next to the module colour panel). Metabolite names in bold were additionally associated with a cognitive outcome in our cognition analysis. Source data are present in Supplementary Table 1 & 3). *AD = Alzheimer’s disease, DMTPA = 2,3 Dihydroxy-5-methylthio-4-pentenoic acid, NSHD = National Survey of Health and Development, PRS = polygenic risk score*

### Polygenic risk scores

We investigated whether modules and hub metabolites that associated with an imaging measure (p_FDR_<0.05) were also associated with polygenic risk for AD (*APOE* region included and excluded). We observed no significant relationships following FDR correction. At the nominal threshold, we report relationships between higher AD PRS and decreased levels of five hub metabolites (Figure 3 and Supplementary Table 3). Three DAG and phosphoethanolamine hub metabolites were associated with the *APOE* AD PRS only (P_T_=5×10-8; ß range=-0.061 to -0.050, p range=0.012 to 0.04, p_FDR_>0.05). Two PUFA were associated with the non-*APOE* AD PRS (P_T_=0.1; DPA: ß =-0.077, 95%CI=[0.13,-0.029], p=0.0018; linolenate: ß =-0.054, 95%CI=[-0.10,-0.0056], p=0.029, p_FDR_>0.05); these associations weakened with the *APOE* region additionally included (P_T_=0.1; DPA: ß =-0.058, 95%CI=[-0.11,-0.0097], p=0.019, linolenate: ß=-0.040, 95%CI=[-0.088,0.0088], p=0.11). No relationships were observed for metabolite modules at either the nominal or adjusted level of significance.

### Additional analysis

Full results from our additional analyses are discussed in Supplementary Notes. Briefly, modules showed moderate to large preservation in the Insight 46 subset of the NSHD (Supplementary Figure 1). Following Bonferroni correction (module: p<3.57×10-3; hub metabolite: p<1.06×10-3) all modules and nine metabolites remained associated with an outcome, with 21 no longer reaching the adjusted level of significance. Further adjustment for life course factors resulted in minimal changes (Supplementary Table 4 & 5).

## Discussion

In a population-based cohort, we identified three modules of coexpressed lipids (phospholipids and DAGs, fatty acids, and sphingolipids) that were associated with whole-brain or hippocampal volume, and 22 highly connected metabolites within these that present as potential markers for additional study. We report no significant metabolic associations for Aβ status following multiple testing correction, nor for AD polygenic risk in our genetic analyses, although some relationships were seen at the nominal level. Taken together, this suggests that these findings may be relevant to later life brain structure but non-specific to AD.

### Fatty acids in whole-brain volume

First, we found that higher expression of the blue module – enriched in several fatty acid pathways – was associated with smaller whole-brain volumes. We identified eight hub metabolites in this module, belonging to fatty acid pathways (long chain PUFA (n3 and n6); monohydroxy; dicarboxylate; medium chain), as well as glycerol from the glycerolipid metabolism pathway. These pathways are tightly linked: fatty acids and glycerol constitute phospholipids and triglycerides, and dicarboxylate and monohydroxy fatty acids are oxidative products of PUFA and other fatty acids.

Upregulation of this module and hub metabolites may thus represent changes in lipid metabolism, including enhanced lipid breakdown, accumulation of free fatty acids and glycerol in the blood, and alterations in fatty acid oxidation – all of which have been linked to neurodegeneration and AD ^31–33^. Worth noting however, is that while n6 PUFA have been typically linked to risk of neurodegenerative disease, n3 have been linked to decreased risk ^34^, although both are an area of contention ^35^. Here, we identified one hub (docosapentaenoate; DPA, 22:5n3) to be an n3 fatty acid, contrasting with the general consensus among the literature. We additionally observed some suggestive evidence of an association between AD genetic risk and PUFA, particularly with the *APOE* region removed; a higher AD PRS was associated with decreased levels of two long chain PUFA hubs (linolenate (18:3n3 or 3n6) and DPA) at the nominal level. These effect directions align with those reported in the literature, but were in the unexpected direction based on our findings in our imaging analysis. Nevertheless, we note caution in interpreting these findings until they are replicated in a larger, independent sample.

We further identified two module hubs (16-hydroxypalmitate and hexadecanedioate) which are products of microsomal omega-oxidation – a minor oxidation pathway for fatty acids. Accumulation thus points to defects in mitochondrial β-oxidation pathways, perhaps induced by an overload of free fatty acids or vice versa ^36,37^. Notably, defective β-oxidation and compensatory omega-oxidation pathways are thought to induce oxidative stress ^38,39^ – a key mechanism linked to neurodegeneration, which is marked among other things by reduction in brain volume ^40^. Nevertheless, to our knowledge, these metabolites have not been linked to brain health and neurodegeneration previously, although they have been found to play a role in other phenotypes, such as mortality and blood pressure ^36^.

### Phospholipids and DAGs in whole-brain volume

Similar to the module of fatty acids, higher expression of the brown module – enriched in PEs and DAGs – associated with smaller whole-brain volumes. These pathways have key roles in membrane structure and cell signalling: PEs are a class of phospholipid which form important components of cellular membranes ^41^, and are a precursor to DAGs, which are components of cellular membranes and secondary messengers ^42^. We similarly reported associations between this module and short-term memory in our previous analysis of cognition in the NSHD; these associations were in the same direction, although they were mostly explained by BMI and lipid medication ^10^. Here, our results were independent of these factors with minimal attenuations overall.

Within the module, we identified four hub metabolites – two phospholipids and two DAGs. Both subclasses have been previously linked to AD and neurodegeneration ^43,44^, and increased serum concentrations have been hypothesised to represent alterations in membrane integrity and subsequent degradation ^45,46^, although some associations in the opposite direction have also been reported ^32,47^. Three module hubs were additionally associated with an AD PRS at the nominal level; this relationship appeared to be driven predominantly by *APOE* and may thus reflect pleiotropic pathways. Again, these were in the opposite direction to our findings for whole-brain volume and did not survive multiple testing correction.

### Sphingolipids in hippocampal volume and whole-brain volume

We highlighted associations between higher levels of sphingolipids, a lipid class that contain important constituents of cellular membranes ^48^, and larger hippocampal and whole-brain volumes. Our findings were observed for sphingomyelins in particular, a subclass that are especially abundant in the CNS, where they form pivotal components of neuronal membranes and play key roles in signal transduction ^49^. Given their biological role, it is unsurprising that sphingolipids have been previously linked to brain health and pathology ^50,51^. Here, we report associations between a module enriched in sphingolipids and hippocampal volume, as well as several sphingomyelin hub metabolites and whole-brain volume.

We previously reported similar findings for sphingolipids and several cognitive outcomes; however, after we adjusted for childhood cognition and social factors – particularly childhood cognition and education – these were entirely attenuated ^10^. Interestingly, we did not observe the same pattern here, with relationships showing negligible attenuations overall, although it is worth noting that childhood cognition and education show much weaker associations with structural brain measures and thus less likely to confound associations. We hypothesised that attenuations could represent earlier relationships we are unable to capture without longitudinal metabolite data, shared genetic or environmental underpinnings, or confounding by reverse causation, i.e. increased sphingolipid levels may be consequential to a higher level of cognition in early life, and education may be capturing shared components. With no earlier measure of brain volume, the latter could extend to our present findings, although previous research has linked sphingomyelins to longitudinal markers of pathology ^50,52,53^. Another possibility is that sphingolipids may have different involvements in cognition and later life brain volume, or may be particularly important during sensitive age periods, for example in cognitive development as well as during vulnerable periods in later life with regard to structural integrity ^48^. Expanding this to longitudinal data, alongside interrogating relationships using MR, will allow for a greater insight into our findings.

### Limited relationships were seen for Aβ status and AD polygenic risk

Interestingly, we saw limited metabolic relationships for Aβ status; no module showed associations (p>0.05) and a handful of metabolites were associated at the nominal threshold only. Possibly there are no robust associations between these metabolites and Aβ, or independent of *APOE*, although other work has reported differently ^5,7^. Alternatively, this may reflect power, particularly given the relatively young age of Insight 46 participants and the fact that they have evidence for Aβ accumulation but not frank dementia. Expanding our analyses to further sweeps will be necessary to address this, by enrolling both a larger sample size and of individuals with detectable Aβ burden, as well as longer term follow-up. Furthermore, five metabolites were associated with an AD PRS at the nominal level, but none survived multiple testing correction and no other metabolic associations were seen at either threshold. As modules and metabolites were selected for PRS analyses based on showing significant associations in our imaging analyses, which were observed for whole-brain or hippocampal volume, and not Aβ, this suggests that these metabolites may not be specific to AD. Alternatively, it may also reflect power; expanding our work in larger samples is warranted.

### Strengths and limitations

Strengths of the study include age-matched cohort with information on a large range of confounders across the life course, including the rarely available measure of childhood cognition, as well as data on both imaging and subdomains of cognition. Moreover, the metabolomics data in this study represent a far more comprehensive proportion of the metabolome than in past clinical metabolomic studies of neuroimaging parameters ^7,53^. Nevertheless, the results of this study should be interpreted in the context of the following limitations. First, our findings may not extend to the general population. Study participants are all white and, compared to the full NSHD cohort, participants enrolled in Insight 46 were on average of slightly better self-rated health, cognitive ability, and SEP. Next, our findings may be subject to residual confounding; further study is needed to disentangle causal relationships. Finally, as there are currently few cohorts with genetic, serum LC-MS, and brain imaging data, we are not yet able to replicate our analyses elsewhere.

## Conclusions

Our findings highlight relationships between groups of lipids and structural brain measures, as well as key metabolites within these that are likely to be driving associations. Future work should be directed towards understanding if these metabolites associate with longitudinal changes in brain volumes, and whether relationships are causal; this could advance our understanding of brain health and neurodegeneration, and reveal possible targets of intervention.

## Data Availability

Anonymised data are available upon request to bona fide researchers (https://skylark.ucl.ac.uk/NSHD/doku.php).

## Declarations

## Consent for publication

Not applicable.

## Authors’ contributions

RG, JMS, MR, and PP designed and set up the research and were responsible for the manuscript. RG performed the primary data analysis and wrote the first draft of the manuscript.. MAS, AW, SNJ, DMS, TDP, CAL, IBM, DMC, CHS, WC, DLT, SK, NCF, JMS, MR, and PP were involved in data acquisition and generation. RG, JL, JX, AH, LG, MR, and PP provided input on data analysis and interpretation. All authors read and approved the final manuscript.

## Acknowledgements

We are very grateful to those study members who helped in the design of the study through focus groups, and to the participants both for their contributions to Insight 46 and for their commitments to research over the last seven decades. We are grateful to the radiographers and nuclear medicine physicians at the UCL Institute of Nuclear Medicine, and to the staff at the Leonard Wolfson Experimental Neurology Centre at UCL. We are particularly indebted to the support of the late Dr Chris Clark of Avid Radiopharmaceuticals who championed this study from its outset.

We thank the International Genomics of Alzheimer’s Project (IGAP) for providing summary results data for these analyses. The investigators within IGAP contributed to the design and implementation of IGAP and/or provided data but did not participate in analysis or writing of this report.

We acknowledge use of the research computing facility at King’s College London, *Rosalind* (https://rosalind.kcl.ac.uk), which is delivered in partnership with the National Institute for Health Research (NIHR) Biomedical Research Centres at South London & Maudsley and Guy’s & St. Thomas’ NHS Foundation Trusts, and part-funded by capital equipment grants from the Maudsley Charity (award 980) and Guy’s & St. Thomas’ Charity (TR130505). We further acknowledge the use of BioRender.com in the creation of Figure 1.

## Abbreviations

Aβ: amyloid-beta
AD: Alzheimer’s disease
BMI: body mass index
DAG: diacylglycerol
FDR: false discovery rate
FLAIR: weighted fluid-attenuated inversion recovery
KEGG: Kyoto Encyclopaedia of Genes and Genomes
kME: module membership
LCMS: liquid chromatography-mass spectrometry
MR: Mendelian randomisation
n3 and n6: Omega 3 and 6
NSHD: National Survey of Health and Development
PC: phosphatidylcholine
PE: phosphatidylethanolamine
PRS: polygenic risk score
PUFA: polyunsaturated fatty acid
QC: quality control
SEP: socioeconomic position
SUVR: standardised uptake volume ratio
WGCNA: weighted gene coexpression network analysis

