## Supplementary figures and images for "Investigating associations between blood metabolites, later life brain imaging measures, and genetic risk for Alzheimer’s disease" (from another version of this preprint: version 2, 2022-02-03)

### Supplementary Figure 1

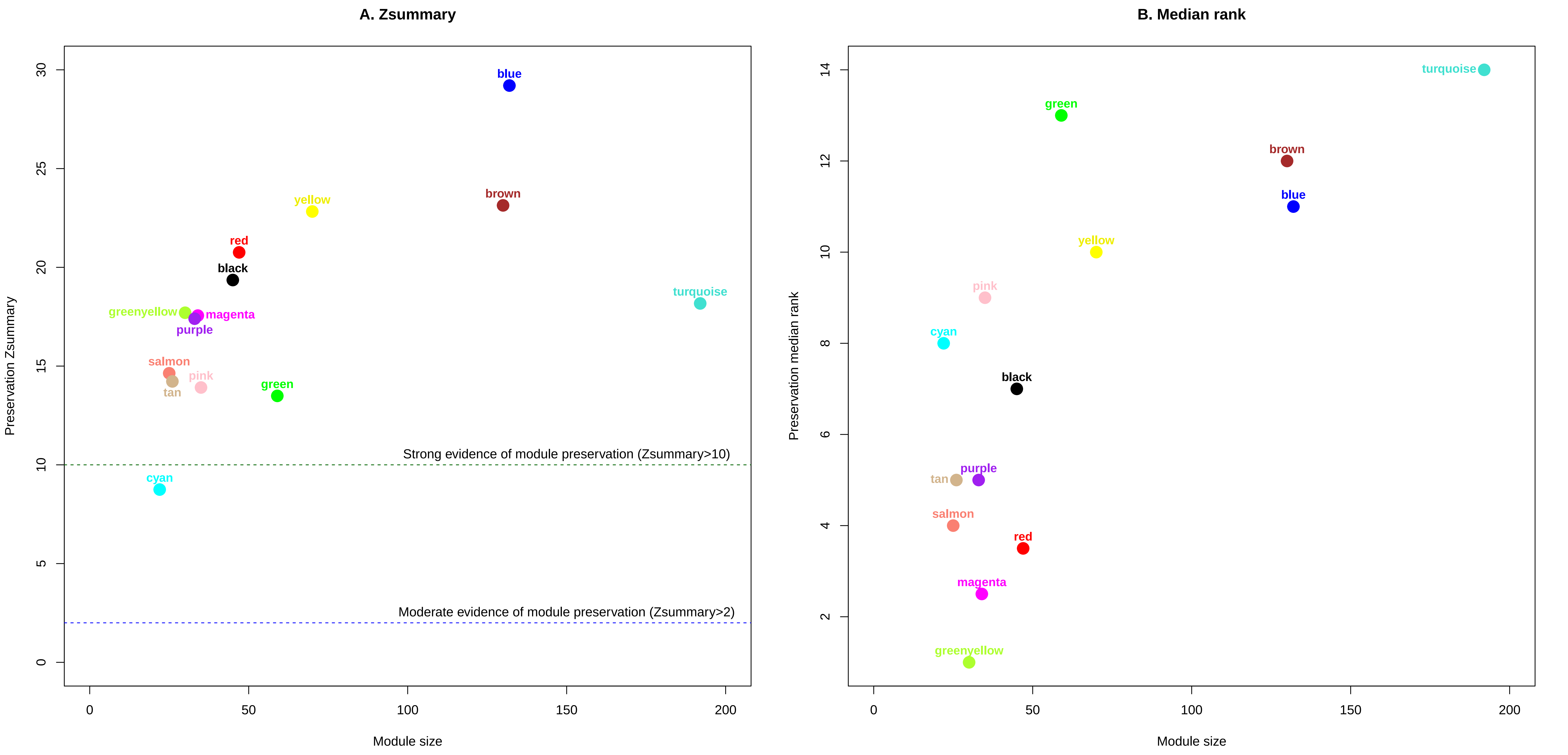
