## Supplementary Notes for "Investigating associations between blood metabolites, later life brain imaging measures, and genetic risk for Alzheimer’s disease" (from another version of this preprint: version 2, 2022-02-03)

### Metabolomic profiling

**Information provided by Metabolon Inc**

**Sample Accessioning:** Following receipt, samples were inventoried and immediately stored at -80^o^C. Each sample received was accessioned into the Metabolon LIMS system and was assigned by the LIMS a unique identifier that was associated with the original source identifier only. This identifier was used to track all sample handling, tasks, results, etc. The samples (and all derived aliquots) were tracked by the LIMS system. All portions of any sample were automatically assigned their own unique identifiers by the LIMS when a new task was created; the relationship of these samples was also tracked. All samples were maintained at -80^o^C until processed.

**Sample Preparation:** Samples were prepared using the automated MicroLab STAR® system from Hamilton Company. Several recovery standards were added prior to the first step in the extraction process for QC purposes. To remove protein, dissociate small molecules bound to protein or trapped in the precipitated protein matrix, and to recover chemically diverse metabolites, proteins were precipitated with methanol under vigorous shaking for 2 min (Glen Mills GenoGrinder 2000) followed by centrifugation. The resulting extract was divided into five fractions: two for analysis by two separate reverse phase (RP)/UPLC-MS/MS methods with positive ion mode electrospray ionization (ESI), one for analysis by RP/UPLC-MS/MS with negative ion mode ESI, one for analysis by HILIC/UPLC-MS/MS with negative ion mode ESI, and one sample was reserved for backup. Samples were placed briefly on a TurboVap® (Zymark) to remove the organic solvent. The sample extracts were stored overnight under nitrogen before preparation for analysis.

**QA/QC:** Several types of controls were analyzed in concert with the experimental samples: a pooled matrix sample generated by taking a small volume of each experimental sample (or alternatively, use of a pool of well-characterized human plasma) served as a technical replicate throughout the data set; extracted water samples served as process blanks; and a cocktail of QC standards that were carefully chosen not to interfere with the measurement of endogenous compounds were spiked into every analyzed sample, allowed instrument performance monitoring and aided chromatographic alignment.Instrument variability was determined by calculating the median relative standard deviation (RSD) for the standards that were added to each sample prior to injection into the mass spectrometers. Overall process variability was determined by calculating the median RSD for all endogenous metabolites (i.e., non-instrument standards) present in 100% of the pooled matrix samples. Experimental samples were randomized across the platform run with QC samples spaced evenly among the injections.

**Ultrahigh Performance Liquid Chromatography-Tandem Mass Spectroscopy (UPLC-MS/MS):** All methods utilized a Waters ACQUITY ultra-performance liquid chromatography (UPLC) and a Thermo Scientific Q-Exactive high resolution/accurate mass spectrometer interfaced with a heated electrospray ionization (HESI-II) source and Orbitrap mass analyzer operated at 35,000 mass resolution. The sample extract was dried then reconstituted in solvents compatible to each of the four methods. Each reconstitution solvent contained a series of standards at fixed concentrations to ensure injection and chromatographic consistency. One aliquot was analyzed using acidic positive ion conditions, chromatographically optimized for more hydrophilic compounds. In this method, the extract was gradient eluted from a C18 column (Waters UPLC BEH C18-2.1x100 mm, 1.7 µm) using water and methanol, containing 0.05% perfluoropentanoic acid (PFPA) and 0.1% formic acid (FA). Another aliquot was also analyzed using acidic positive ion conditions, however it was chromatographically optimized for more hydrophobic compounds. In this method, the extract was gradient eluted from the same afore mentioned C18 column using methanol, acetonitrile, water, 0.05% PFPA and 0.01% FA and was operated at an overall higher organic content. Another aliquot was analyzed using basic negative ion optimized conditions using a separate dedicated C18 column. The basic extracts were gradient eluted from the column using methanol and water, however with 6.5mM Ammonium Bicarbonate at pH 8. The fourth aliquot was analyzed via negative ionization following elution from a HILIC column (Waters UPLC BEH Amide 2.1x150 mm, 1.7 µm) using a gradient consisting of water and acetonitrile with 10mM Ammonium Formate, pH 10.8. The MS analysis alternated between MS and data-dependent MS^n^ scans using dynamic exclusion. The scan range varied slighted between methods but covered 70-1000 m/z. Raw data files are archived and extracted as described below.

**Data Extraction and Compound Identification:** Raw data was extracted, peak-identified and QC processed using Metabolon’s hardware and software. These systems are built on a web-service platform utilizing Microsoft’s .NET technologies, which run on high-performance application servers and fiber-channel storage arrays in clusters to provide active failover and load-balancing. Compounds were identified by comparison to library entries of purified standards or recurrent unknown entities. Metabolon maintains a library based on authenticated standards that contains the retention time/index (RI), mass to charge ratio (*m/z)*, and chromatographic data (including MS/MS spectral data) on all molecules present in the library. Furthermore, biochemical identifications are based on three criteria: retention index within a narrow RI window of the proposed identification, accurate mass match to the library +/- 10 ppm, and the MS/MS forward and reverse scores between the experimental data and authentic standards. The MS/MS scores are based on a comparison of the ions present in the experimental spectrum to the ions present in the library spectrum. While there may be similarities between these molecules based on one of these factors, the use of all three data points can be utilized to distinguish and differentiate biochemicals. More than 3300 commercially available purified standard compounds have been acquired and registered into LIMS for analysis on all platforms for determination of their analytical characteristics. Additional mass spectral entries have been created for structurally unnamed biochemicals, which have been identified by virtue of their recurrent nature (both chromatographic and mass spectral). These compounds have the potential to be identified by future acquisition of a matching purified standard or by classical structural analysis.

**Curation:** A variety of curation procedures were carried out to ensure that a high quality data set was made available for statistical analysis and data interpretation. The QC and curation processes were designed to ensure accurate and consistent identification of true chemical entities, and to remove those representing system artifacts, mis-assignments, and background noise. Metabolon data analysts use proprietary visualization and interpretation software to confirm the consistency of peak identification among the various samples. Library matches for each compound were checked for each sample and corrected if necessary.

**Metabolite Quantification and Data Normalization:** Peaks were quantified using area-under-the-curve. A data normalization step was performed to correct variation resulting from instrument inter-day tuning differences. Essentially, each compound was corrected in run-day blocks by registering the medians to equal one (1.00) and normalizing each data point proportionately.

### Metabolite QC

The figure below details metabolomics quality control (QC) steps and resulting numbers of metabolites, participants, and participants with available outcome data in this study.

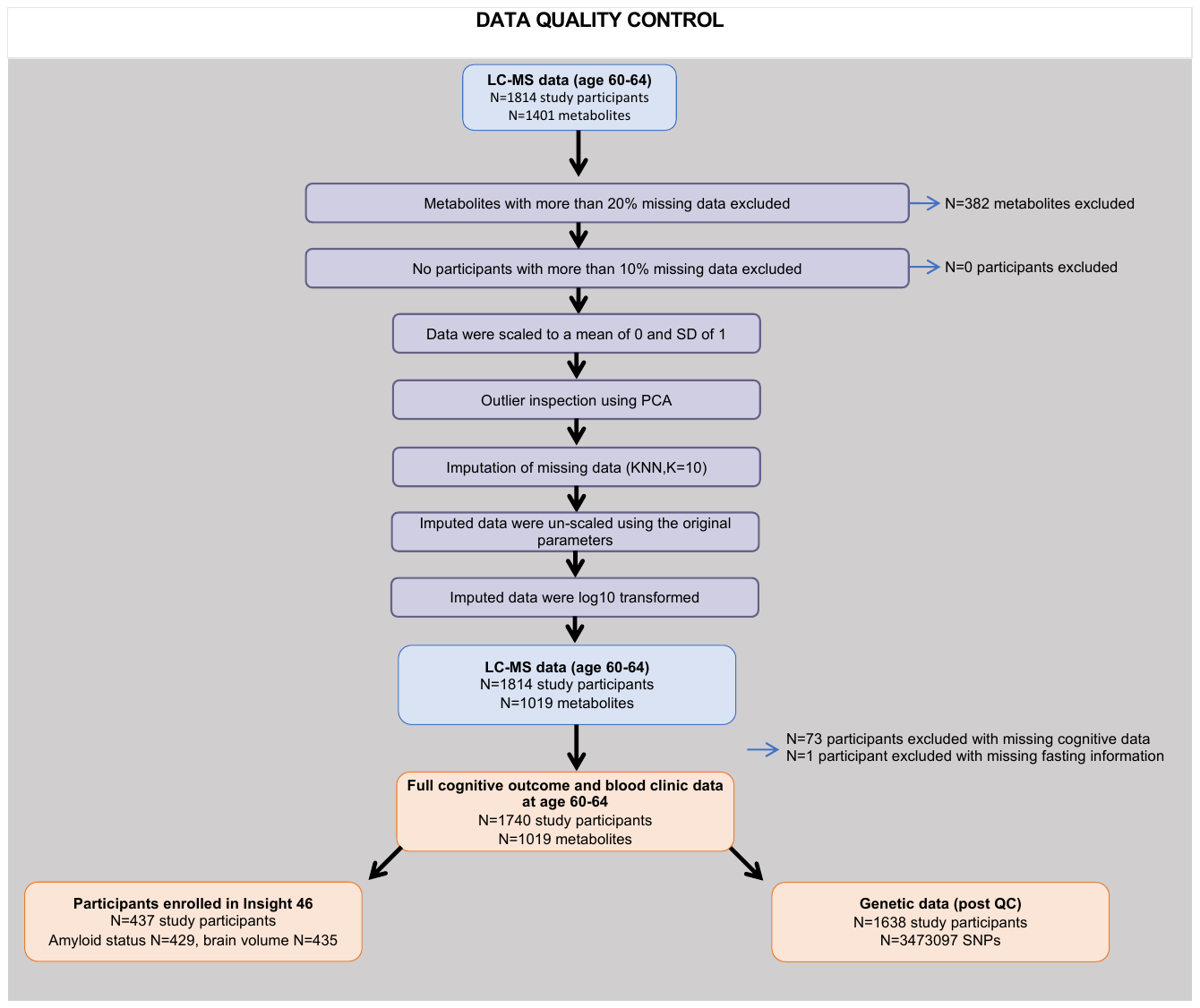

### Genetic QC

Genetic data were extracted from blood samples collected at ages 53 and 60-64 and assayed using two genotyping chips (NeuroX2, Infinium NeuroConsortium Array, Illumina, inc; and DrugDev, Illumina, inc.). The initial QC and imputation were performed centrally by the National Survey of Health and Development (NSHD) study team, using a similar pipeline to (1). Prior to imputation, non-autosomal SNPs were removed, sex checks were performed, and relatedness was checked. Chips were merged and cleaned using a tool by W. Rayner (<https://www.well.ox.ac.uk/~wrayner/tools/>), which performs checks for strand consistency, allele names, position, Ref/Alt assignments, and minor allele frequency (MAF) differences with the Haplotype Reference Consortium (HRC) reference panel (set to >0.1 for the NSHD) (1). Data were imputed using the Sanger Imputation Server (<https://imputation.sanger.ac.uk/>) using the HRC v1.1 as a reference panel (2), EAGLE2 for phasing (3) and PBWT for imputation (4). Following imputation, multiallelic variants and variants with an INFO score of <0.7 were removed, and imputed genotypes with a posterior probability of <0.9 were set to missing. Variants with a call rate of <90%, MAF <1x10^-5^ or Hardy-Weinberg p-value <5x10^-7^ were also removed. Genetic ancestry was inferred using SNPweights (5) and projected onto the HapMap 3 reference panel (6). Ten principal components were derived.

For the purpose of this study, we additionally removed rare variants (MAF <5%), as well as variants with a low call rate (<98%) or deviating from Hardy-Weinberg equilibrium (1x10^-5^). Participants with a low call rate (<98%), mismatching sex, or that were related (PIHAT <0.1) were also removed. All QC was performed using PLINK v1.9 (https://www.cog-genomics.org/plink2). Following QC, genetic and metabolomic data were available for 1638 participants.

### Polygenic scores

SNPs included in the APOE and non-APOE polygenic risk scores, at the 5x10^-8^ p-value threshold, can be found below.

All SNPs were included for the APOE PRS. SNPs in bold were included for the non-APOE PRS.

| **Chromosome** | **SNP** | **Base pair** | **P-value in summary statistics** |
| --- | --- | --- | --- |
| 1 | **rs679515** | 207750568 | 1.6e-16 |
| 2 | **rs6733839** | 127892810 | 4.0e-28 |
| 6 | **rs9381563** | 47432637 | 2.9e-08 |
| 7 | **rs11767557** | 143109139 | 1.6e-08 |
| 8 | **rs73223431** | 27219987 | 8.3e-10 |
| 8 | **rs1532276** | 27466157 | 1.7e-16 |
| 11 | **rs67472071** | 47391745 | 1.1e-10 |
| 11 | **rs10838738** | 47663049 | 6.0e-09 |
| 11 | **rs1582763** | 60021948 | 1.2e-16 |
| 11 | **rs3851179** | 85868640 | 5.8e-16 |
| 19 | **rs12151021** | 1050874 | 2.6e-10 |
| 19 | rs56261258 | 45149613 | 2.8e-15 |
| 19 | rs429358 | 45411941 | 1.2 × 10^−881^ |
| 19 | rs7257916 | 45482884 | 4.5e-10 |
| 19 | rs59839536 | 45689977 | 3.0e-21 |

### Additional analyses

#### Module preservation

As modules were curated in the NSHD, we investigated whether these were preserved in the Insight 46 subset. Module preservation was measured using the Zsummary score from the modulePreservation function (200 permutations with the NSHD as the reference dataset) within the WGCNA package; this score is an aggregated measure of preservation statistics measuring similarity in module density and connectivity patterns. A Zsummary of >10 has been suggested to indicate strong evidence of module preservation, Zsummary>2 moderate preservation, and Zsummary <2 no module preservation (9). Module preservation analyses indicated that metabolite modules were preserved in Insight 46, with all modules showing moderate to large evidence of preservation (Zsummary range=8.75 to 29.2, Supplementary Notes & Supplementary Figure 1).

#### Bonferroni correction

In addition to false discovery rate correction, we applied a more conservative Bonferroni correction, adjusting for the number of independent metabolites and modules. For module relationships, a Bonferroni-adjusted significance threshold was computed as: 0.05/14 modules, p<3.57x10^-3^. As metabolites are highly correlated, and hub metabolites residing in the same module particularly so by definition, we adjusted for the number of principal components (PCs) explaining >95% variance in all 116 hubs investigated: 0.05/47 PCs, p<1.06x10^-3^. This approach has been applied elsewhere (8,10). Following Bonferroni correction, all modules remained associated with an outcome. However, the brown module became nominally significant in model 3. Nine metabolites remained at the adjusted threshold (blue: 16-hydroxypalmitate; docosapentaneoate (22:5n3)*; yellow: sphingomyelin (d18:2/23:1)*, sphingomyelin (d18:2/16:0, d18:1/16:1)*, sphingomyelin (d18:1/22:2, d18:2/22:1, d16:1/24.2)*, sphingomyelin (d17:2/16:0, d18:2/15:0)*, sphingomyelin (d18:2/21:0, d16:2/23:0)*, sphingomyelin (d18:1/25:0, d19:0/24:1, d20:1/23:0, d19:1/24:0)*, sphingomyelin (d18:1/19:0, d19:1/24:0)*), and 21 became nominally significant. No changes were seen for polygenic risk score analyses.

#### Lifestyle analyses

We explored associations between lifestyle covariables, imaging outcomes, and significant (pFDR<0.05) modules and metabolites using regression analyses. Lifestyle covariables included the following: lifetime smoking (pack years between 20 and 60-64y), alcohol intake (none, light-to-moderate, heavy; 3-5 day diet diaries at 36, 43, 53 and 60-64y), systolic blood pressure (second measurement; 60-64y), physical activity (none, 1-4 times per week, >4 times per week during month prior; 60-64y), diet (sex-specific quintiles reflecting adherence to the Dietary Approaches to Stop Hypertension (DASH) diet (7); diet diaries 60-64y). Further details can be found in (8). In the case of any significant (p<0.05) associations, we repeated model 3 analyses with these factors additionally included. We identified relationships between smoking and alcohol consumption and whole-brain volume (Supplementary Table 4). Following additional adjustment for these variables in model 3, all significant (pFDR<0.05) metabolites and modules remained at the adjusted or nominal threshold, with the exception of glycerol (Supplementary Table 5).

### Software & packages

1. RStudio version 3.6.0

Package Version

arsenal	3.6.3

cowplot	1.1.1

data.table 1.12.8

dplyr	1.0.7

foreign 0.8-71

ggdendro 0.1-20

ggh4x 0.1.2.1

ggplot2	3.3.3

ggpubr	0.3.0

ggtext	0.1.1

gtable	0.3.0

Hmisc	4.5-0

kableExtra	1.3.4

knitr	1.30

lubridate 1.7.10

magrittr	2.0.1

mice 3.11.0

openxlsx	4.2.3

plyr	1.8.6

readstata13	0.9.2

tableone 0.11.1

tidyr	1.1.2

2. PLINK version 1.9 (<https://www.cog-genomics.org/plink2>) (11)
3. PRSice version 2 (<https://www.prsice.info/>) (12)

### Participant characteristics split by amyloid status

|  |  | Participants with imaging and metabolite data | | | |
| --- | --- | --- | --- | --- | --- |
|  |  | Overall | Amyloid negative | Amyloid positive | Missing (%) |
| n |  | 437 | 348 | 81 |  |
| Sex, N (%) | Male | 229 (52.4) | 178 (51.1) | 45 (55.6) | 0 |
|  | Female | 208 (47.6) | 170 (48.9) | 36 (44.4) |  |
| Age at scan (years), mean (SD) |  | 70.7 (0.7) | 70.7 (0.7) | 70.6 (0.6) | 0 |
| APOE4 carrier, N (%) | Non-carrier | 306 (70.3) | 269 (77.7) | 35 (43.2) | 0.5 |
|  | Carrier | 129 (29.7) | 77 (22.3) | 46 (56.8) |  |
| Hippocampal volume in ml, mean (SD) |  | 3.1 (0.3) | 3.1 (0.3) | 3.1 (0.3) | 0.5 |
| Brain volume in ml, mean (SD) |  | 1101.8 (99.3) | 1095.5 (98.1) | 1120.9 (100.5) | 0.5 |
| Total intracranial volume in ml, mean (SD) |  | 1435.9 (132.2) | 1428.3 (132.6) | 1457.9 (125.3) | 0.5 |
| Age at blood collection (years), mean (SD) |  | 63.3 (1.1) | 63.3 (1.1) | 63.3 (1.0) | 0 |
| Fasting at blood collection , N (%) | No | 6 (1.4) | 5 (1.4) | 0 (0.0) | 0 |
|  | Yes | 431 (98.6) | 343 (98.6) | 81 (100.0) |  |
| Lipid medication use (age 60-64), N (%) | No | 335 (76.7) | 273 (78.4) | 57 (70.4) | 0 |
|  | Yes | 102 (23.3) | 75 (21.6) | 24 (29.6) |  |
| Body mass index (age 60-64) in kg/m2, mean (SD) |  | 27.4 (4.0) | 27.5 (4.0) | 26.9 (3.8) | 0 |
| Childhood cognitive ability (age 15)*, z-score, mean (SD) |  | 0.5 (0.7) | 0.5 (0.7) | 0.5 (0.7) | 8.2 |
| Childhood socioeconomic position (age 11), N (%) | Unskilled | 18 (4.1) | 13 (3.7) | 4 (5.0) | 0.5 |
|  | Partly skilled | 64 (14.7) | 58 (16.7) | 6 (7.5) |  |
|  | Manual skilled | 106 (24.4) | 79 (22.8) | 23 (28.7) |  |
|  | Nonmanual skilled | 90 (20.7) | 71 (20.5) | 18 (22.5) |  |
|  | Intermediate | 110 (25.3) | 88 (25.4) | 20 (25.0) |  |
|  | Professional | 47 (10.8) | 38 (11.0) | 9 (11.2) |  |
| Adult socioeconomic position (age 53), N (%) | Unskilled | 4 (0.9) | 3 (0.9) | 1 (1.2) | 0 |
|  | Partly skilled | 22 (5.0) | 19 (5.5) | 3 (3.7) |  |
|  | Manual skilled | 40 (9.2) | 32 (9.2) | 7 (8.6) |  |
|  | Nonmanual skilled | 91 (20.8) | 78 (22.4) | 13 (16.0) |  |
|  | Intermediate | 225 (51.5) | 176 (50.6) | 45 (55.6) |  |
|  | Professional | 55 (12.6) | 40 (11.5) | 12 (14.8) |  |
| Highest educational attainment (age 26), N (%) | No qualification | 65 (15.3) | 51 (15.2) | 12 (14.8) | 3 |
|  | Up to GCSE | 125 (29.5) | 99 (29.6) | 25 (30.9) |  |
|  | A-level or higher | 234 (55.2) | 185 (55.2) | 44 (54.3) |  |

*Childhood cognition Z-scores were calculated in the full National Survey of Health and Development cohort (N=5362).
